# Infant malnutrition in low- and middle-income countries: assessment and prevalence of small and nutritionally at-risk infants aged under 6 months in 54 Demographic & Health Survey datasets

**DOI:** 10.1101/2021.12.23.21268306

**Authors:** Marko Kerac, Philip T James, Marie G McGrath, Eilise Brennan, Charles Opondo, Severine Frison

**Author notes:** Corresponding author: Philip T James, Emergency Nutrition Network, Oxford, UK.

## Abstract

**Background:** There is increasing global focus on malnutrition in infants aged under 6 months (u6m) but evidence on how best to identify and manage at-risk individuals is sparse. Our objectives were to: explore data quality of commonly used anthropometric indicators; describe prevalence and disease burden of infant u6m malnutrition; compare wasting and underweight as measures of malnutrition by determining the strength and consistency of associations with biologically plausible risk factors.

**Methods:** We performed a cross-sectional secondary analysis of Demographic and Health Survey (DHS) datasets, focussing on infants u6m. We calculated underweight (low weight-for-age), wasting (low weight-for-length), stunting (low length-for-age), and concurrent wasting and stunting. We explored data quality by recording extreme (flagged, as per standard criteria) or missing values. We calculated the population-weighted prevalence of each type of malnutrition and extrapolated the burden to all low- and middle-income countries (LMICs). We explored associations between infant, maternal and household risk factors with underweight and wasting using logistic regression models.

**Results:** We analysed 54 DHS surveys. Data quality in terms of refusals and missingness was similar for both weight and length. There were more extreme (flagged) values for length-based measures (6.1% flagged for weight-for-length, 4.8% for length-for-age) than for weight-for-age (1.0% flagged). Overall, 20.1% of infants (95% CI: 19.5, 20.7) were underweight, 21.3% (95% CI: 20.7, 22.3) were wasted, 17.6% (95% CI: 17.0, 18.2) were stunted, and 2.0% (95% CI: 1.8, 2.2) were concurrently wasted and stunted. This corresponds to an estimated burden in LMICs of 23.8m underweight infants, 24.5m wasted infants, 21.5m stunted infants and 2.2m concurrently wasted and stunted. Logistic regression models showed that numerous risk factors were associated with wasting and underweight. Effect sizes of risk factors tended to be stronger and more consistently associated with underweight compared to wasting.

**Conclusion:** Malnutrition in infants u6m is a major problem in LMICs. This is true whether assessed by underweight, wasting or stunting. Our data build on other evidence suggesting that underweight may be a better anthropometric case definition than wasting: data quality is better when length is not involved; biologically plausible risk factors are better reflected by an infant being underweight. Future research, ideally from intervention trials, should further explore how best to identify malnourished (small and nutritionally at-risk) infants u6m. For now, treatment programmes should note that many factors might underlie problems in this age group: services should thus consider how to address maternal health and wider social circumstances as well as caring for infants themselves.

## INTRODUCTION

Malnutrition in infants aged under 6 months (u6m) is increasingly recognised as an important global health issue (1-3). Infants are not only at high risk of death in the short-term (4), but risk long term sequelae including adult overweight/obesity (5) and non-communicable disease (6).

Much focus to date has been on infants u6m who are wasted, those with low weight-for-length, one of several ways of defining severe malnutrition (7). 2011 estimates suggest that in low- and middle-income countries (LMICs), some 3.8 million infants u6m were severely wasted, and a further 4.7 million moderately wasted (8). The World Health Organization (WHO) 2013 Guidelines on Severe Acute Malnutrition used severe wasting, weight-for-length <-3 z-scores (standard deviations from population median), as the main admission criterion to treatment programmes (1). Though many of the recommendations in these WHO guidelines were ‘strong’, the underlying evidence-base was recognised as being sparse and overall low quality. Many research and programmatic gaps remain (9, 10).

The WHO is currently updating its severe malnutrition guidelines (11). Infants u6m are among the key groups of interest. Also being explored is how best to identify those most at-risk of adverse mortality/morbidity outcomes. Anthropometric deficit is a widely used, useful but imperfect measure of malnutrition, malnutrition being “any condition in which deficiency, excess or imbalance of energy, protein or other nutrients…adversely affects body function and/or clinical outcome” (12). No single anthropometric measure is a ‘gold standard’ and all have strengths as well as limitations (7). As well as being in the 2013 WHO guidelines, low weight-for-length is also the measure used to define at-risk cases by most current national malnutrition guidelines (13). However, for infants u6m, there are increasing concerns that this:

- Is time-consuming, requiring too much equipment and hence not being practical at-scale (14);
- Is subject to more measurement errors than other anthropometric indicators of malnutrition (15, 16);
- Is poor at identifying those infants at highest risk of death, the key adverse outcome (17, 18).

To contribute to global efforts improving the identification and care of small and nutritionally at-risk infants u6m, our objectives in this paper are to:

1. Explore the quality of data of the three key anthropometric indicators used to define infant malnutrition (weight-for-length, weight-for-age, and length-for-age).
2. Describe the prevalence of wasted, underweight, stunted, and concurrently wasted and stunted infants < 6m overall and by region.
3. Provide an updated estimate of the burden of wasted, underweight, stunted, and concurrently wasted and stunted infants <6m in all LMICs.
4. Compare wasting and underweight as measures of malnutrition by determining strength and consistency of associations with established, biologically plausible risk factors.

## METHODS

### Study design and setting

We performed a cross-sectional secondary analysis of Demographic and Health Survey (DHS) datasets. DHS are “nationally-representative household surveys that provide data for a wide range of monitoring and impact evaluation indicators in the areas of population, health, and nutrition” (19). They are conducted in LMICs and are usually updated in 5-yearly cycles. To facilitate cross-country comparisons and analyses they follow a common, standardized methodology which includes two stage cluster sampling and questionnaires focusing on women, children and household characteristics (20). Raw survey data is free to download after registration on the DHS website (21). Our methodology was informed by a previous paper that performed a risk factor analysis of wasting in infants u6m in 20 countries (22).

### Participant inclusion criteria and sample size

We focused our analyses on infants u6m. Inclusion criteria for the DHS surveys included: the latest survey from a country; conducted in the last 10 years; includes data on the sex, age, weight and length of infants u6m. Surveys were excluded if they were conducted more than 10 years prior to analysis or if key anthropometric data on infants were not collected. Our sample size was constrained by the available DHS datasets fulfilling the above criteria and by the number of infants u6m in each database.

### Database setup and cleaning

We merged individual country files into one large dataset. We generated z-scores as per standard methodology for the WHO’s Child Growth Standards (23), using the WHO Anthro macro for Stata. We explored the quality of data in each of the indicators, recording how many extreme or missing values there were for each. Following standard survey procedures (24), extreme anthropometric values were excluded according to WHO recommendations: length-for-age z-score (LAZ; <-6, >+6), weight-for-length z-score (WLZ; <-5, >+5) and weight-for-age z-score (WAZ; <-6, >+5) (25).

### Anthropometric variables

For the overall survey prevalence estimates we had four main anthropometric indicators:

- wasted infants, defined as WLZ <-2;
- underweight infants, defined as WAZ <-2;
- stunted infants, defined as LAZ <-2;
- infants who were concurrently wasted and stunted, defined as those individuals with WLZ <-2 and HAZ <-2.

For the first three outcomes we defined a severe deficit as <-3 z-scores from the population median, a moderate deficit from <-2 to ≥-3 z-scores, and a ‘normal’ nutritional status for those with ≥-2 z-scores. To generate the survey weights we used population estimates from the online database of the 2019 Revision of World Population Prospects from the United Nations Department of Economic and Social Affairs (26). Population estimates are given for all those aged zero to one year. We used these population estimates for each country at the closest time to the survey year to calculate the appropriate survey weights. We calculated the population-weighted prevalence and 95% confidence intervals of infants stunted, wasted, underweight, and concurrently wasted and stunted. We summarised the outcomes by country, region and overall.

### Total LMIC burden estimates

To estimate the current total LMIC burden of infant undernutrition, we used the same United Nations population database (26) but instead of using population estimates closest to the survey year we used the most up-to-date projections available, those projected for 2020. We used the figures for children aged less than one year, which we halved to estimate the projected population of infants <6 months in each country. We used these figures to calculate an updated population weighting for each country. We then used the pooled, weighted prevalence estimates of our four anthropometric indicators to extrapolate the burden to all LMICs, as defined by World Bank income group categories. We assumed that our 54-country database was representative of all LMICs.

### Risk factor variables associated with underweight and wasted infants

We explored risk factors associated with underweight and wasted infants based on a previous risk-factor paper using a smaller number of DHS datasets (22). We focused on these as they are the two anthropometric criteria most likely to be used in malnutrition treatment programmes given associations with short term risk of mortality (18). The rationale for this analysis was twofold: i) identify malnutrition risk factors in a larger dataset; and ii) explore the strength and consistency of associations with biologically plausible, known risk factors to add to discussions about whether wasting or underweight should be used programmatically to best identify ‘severe malnutrition’ at this age (7).

Variables considered were broadly classed in three categories: infant, maternal and household characteristics. Infant characteristics included age, sex, birth order, birth spacing, place of delivery, delivery by caesarian section (C-section), size at birth, receiving postnatal care (PNC), time of breastfeeding initiation, given anything but breast milk during the first 3 days, being ever breastfed, being currently breastfed, having been exclusively / predominantly breastfed or bottle fed, having a vaccination card, having a BCG vaccination, having timely vaccination, and being ill in the last two weeks (cough, fever, diarrhoea). Maternal characteristics included age, education, BMI, height, input into health decisions, whether 4 or more antenatal care (ANC) visits were received, whether working or not, whether married, and history of previous child deaths. Household characteristics included residence type (urban or rural), water source, time to fetch water, toilet type, and wealth index quintiles. All variables were captured according to standard DHS questionnaire methodology. Details on how questions are asked and how variables were processed are described in full in the official DHS manual (20).

We explored the association of each risk factor with underweight and wasted infants using logistic regression models, adjusted for infant sex, maternal age, marital status, wealth index and mother’s education. In these models we compared those infants who were underweight or wasted with those who had normal status for the anthropometric outcome of interest. All analyses were performed in Stata version 15.0 (*StataCorp* LLC, College Station, TX). We used appropriate survey weighting techniques with Stata’s svyset command.

### Ethics

Ethical approval for this secondary analysis was granted by the London School of Hygiene & Tropical Medicine’s Research Ethics Committee (reference 21896).

## RESULTS

Ninety-three DHS datasets were considered, of which 54 met the inclusion criteria. These comprised 16 surveys from West & Central Africa, 16 from East & Southern Africa, 7 from the Americas, 7 from Southeast Asia & Western Pacific, 4 from Europe and 4 from the Eastern Mediterranean. All surveys were from DHS phases 5-7. From the pooled dataset, a total of 75,804 infants u6m were available for analyses. **Supplementary Table 1** provides the region, country, DHS phase and sample size of each of the included DHS surveys.

### Data quality

**Table 1** shows the descriptive and missing data for the pooled survey dataset. Overall, 49% of the sample infants u6m were female. 1.2% of the caretakers refused to allow their infants’ weight to be measured and 1.8% refused length measurement. Refusals were higher in younger age groups and again were greater for length measurement. Of those who agreed to continue with anthropometric measurements, approximately one quarter of infants then had either a missing weight or a missing length measurement. The proportion of missing measurements was highest for neonates in the first month of life. Of those infants with relevant anthropometric information recorded, 1.0% of WAZ, 6.1% of WLZ and 4.8% of LAZ records were flagged by WHO cleaning criteria as being extreme values and were therefore excluded.

**Table 1:** Descriptive and missing data for anthropometric measurements, by infant age category

|  |  | <1month | 1 month | 2 to <4 months | 4 to <6 months | Total |
| --- | --- | --- | --- | --- | --- | --- |
| Age in months | N (%) | 7315 (9.7) | 13080 (17.3) | 27559 (36.4) | 27850 (36.7) | 75804 (100) |
| Sex (female) | N (%) | 3574 (48.9) | 6339 (48.5) | 13447 (48.8) | 13591 (48.8) | 36951 (48.7) |
| Weight (kg) | Mean (SD) | 3.4 (0.7) | 4.1 (1.0) | 5.2 (1.1) | 6.3 (1.2) | 5.3(1.5) |
| Refused | N (%) | 202 (2.8) | 202 (1.5) | 299 (1.1) | 226 (0.8) | 929 (1.2) |
| Missing | N (%) | 2246 (30.7) | 3250 (24.8) | 6525 (23.7) | 6490 (23.3) | 18511 (24.4) |
| Length (cm) | Mean (SD) | 50.6 (3.9) | 53.6 (4.2) | 58.6 (4.4) | 63.1 (4.2) | 58.8 (6.0) |
| Refused | N (%) | 314 (4.3) | 300 (2.3) | 425 (1.5) | 335 (1.2) | 1374 (1.8) |
| Missing | N (%) | 2430 (33.2) | 3406 (26.0) | 6742 (24.5) | 6650 (23.9) | 19228 (25.4) |
| WHO Flags* |  |  |  |  |  |  |
| WAZ | N (%) | 118 (2.3) | 119 (1.2) | 170 (0.8) | 152 (0.7) | 559 (1.0) |
| WLZ | N (%) | 360 (8.1) | 734 (7.9) | 1256 (6.1) | 1042 (4.9) | 3392 (6.1) |
| LAZ | N (%) | 325 (6.7) | 547 (5.7) | 882 (4.2) | 941 (4.4) | 2695 (4.8) |

\*Excluded data as due to standard survey cleaning criteria: length-for-age z-score (LAZ; <-6, >+6), weight-for-length z-score (WLZ; <-5, >+5) and weight-for-age z-score (WAZ; <-6, >+5)

### Overall prevalence of infants with anthropometric deficits

After excluding refusals, missing records and outliers, 56,734 (74.8%) of the original sample were available for calculation of underweight infants, 51,913 (68.5%) for calculation of wasted infants and 53,881 (71.1%) for calculation of stunted infants (**Table 2**). The overall dataset had a mean (SD) WAZ, WLZ and LAZ just below the reference median (WAZ: -0.75 (1.61), WLZ: -0.58 (1.83), LAZ: -0.27 (2.03)). One fifth of the pooled dataset were underweight, with 8.1% (95% CI: 7.8, 8.5) severely underweight. Similarly, approximately one fifth were wasted, with 9.6% (95% CI: 9.2, 10.0) of those being severely wasted. The proportion of those stunted was very slightly smaller (17.6%), but 8.4% (95% CI: 8.0, 8.8) of these were severely stunted. The proportion of infants concurrently wasted and stunted was 2.0% (95% CI: 1.8, 2.2). In **Supplementary File 1** we present the anthropometric summaries by country and region. Southeast Asia and Western Pacific region had the highest proportion of underweight and wasted infants, respectively (27.3%; 95% CI: 26.4, 28.3 and 28.9%; 95% CI: 27.9,30.0). The Eastern Mediterranean region had the highest proportion of stunted infants (22.2%; 95% CI: 19.7, 24.9).

**Table 2:** Overall population-weighted prevalence* of underweight, wasted, stunted and concurrently wasted and stunted infants < 6m in 54 DHS surveys

|  |  |  |
| --- | --- | --- |
| **Underweight (N=56,734)** |  |  |
| Mean WAZ | Mean (SD) | -0.75 (1.61) |
| Underweight | % (95% CI) | 20.1 (19.5, 20.7) |
| Moderate underweight | % (95% CI) | 11.9 (11.5, 12.4) |
| Severe underweight | % (95% CI) | 8.1 (7.8, 8.5) |
| **Wasting (N=51,913)** |  |  |
| Mean WLZ | Mean (SD) | -0.58 (1.83) |
| Wasted | % (95% CI) | 21.3 (20.7, 22.3) |
| Moderately wasted | % (95% CI) | 11.7 (11.2, 12.3) |
| Severely wasted | % (95% CI) | 9.6 (9.2, 10.0) |
| **Stunting (LAZ) (N=53,881)** |  |  |
| Mean LAZ | Mean (SD) | -0.27 (2.03) |
| Stunted | % (95% CI) | 17.6 (17.0, 18.2) |
| Moderately stunted | % (95% CI) | 9.2 (8.8, 9.7) |
| Severely stunted | % (95% CI) | 8.4 (8.0, 8.8) |
| **Wasted and Stunted (N=50,908)** | % (95% CI) | 2.0 (1.8, 2.2) |

\*Population weighting using UN population database estimates closest to the survey year for each country

### Total LMIC burden estimates

We present the estimated burden of undernutrition extrapolated to all LMICs in **Table 3**, alongside the pooled, population-weighted prevalence estimates updated to 2020 population forecasts. We estimate an approximate total of 23.8m underweight infants, of which 9.5m are severely underweight; 24.5m wasted infants, of which 11.0m are severely wasted; 21.5m stunted infants, of which 10.4m are severely stunted, and 2.2m concurrently wasted and stunted. In **Supplementary File 2** we additionally provide the country-specific burden and the population-weighted prevalence of each type of undernutrition.

**Table 3:**
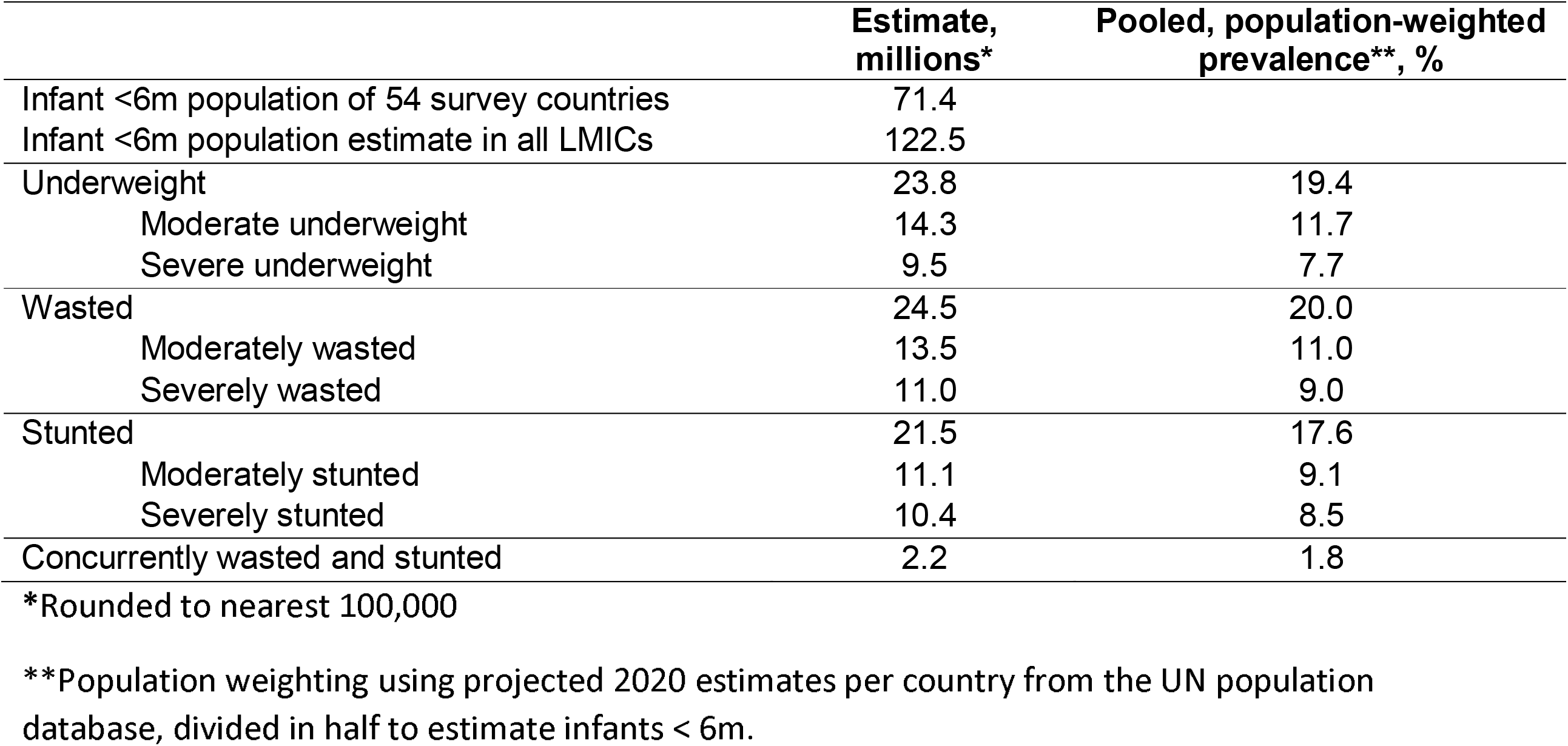
Estimated burden of infant u6m undernutrition in all LMICs.

### Risk factors for underweight and wasted infants

**Table 4** shows the associations between various risk factors and a) underweight and b) wasted infants, using logistic regression models adjusted for sex, age, wealth, mother’s education and marital status. Several risk factors were associated both with decreased odds of being underweight and decreased odds of being wasted (‘protective’ factors). These included having an improved water source, a non-improved toilet compared to an improved one, collecting water (regardless of time taken) compared to having water onsite, maternal age of > 35 years compared to being < 20 years, maternal overweight or obesity compared to normal BMI, mothers having primary education compared to none, mothers who were working versus not working, being large at birth versus normal size, not being the first born, having four or more ANC visits versus none, having a vaccination card, having a BCG vaccine, timely vaccines (DTP and polio), and having a cough in the last two weeks. The final key finding in these analyses was a tendency for effect sizes to be stronger (i.e., larger OR) with underweight compared to wasted infants. For example, the association with wealth index for underweight infants ranges from an OR of 1.44 (poorest, most at-risk) to 0.87 (wealthiest, least at-risk), with two of four categories statistically significant. In contrast, the same association for wasted infants ranges from OR 1.19 to 0.91 with only one statistically significant association. Other risk factor variables where the strength of association is stronger and clearer for underweight infants compared to wasted infants include:

**Table 4:**
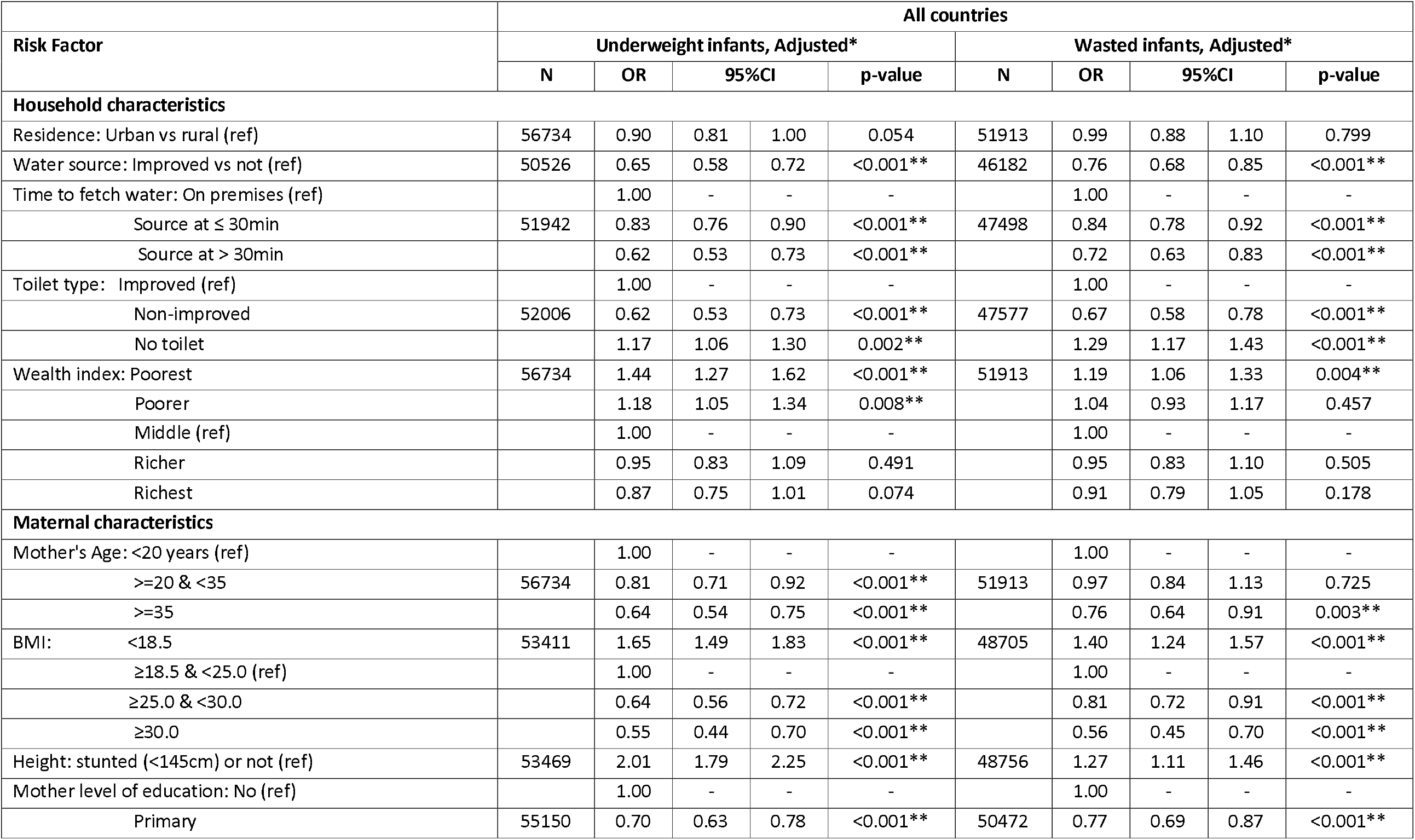
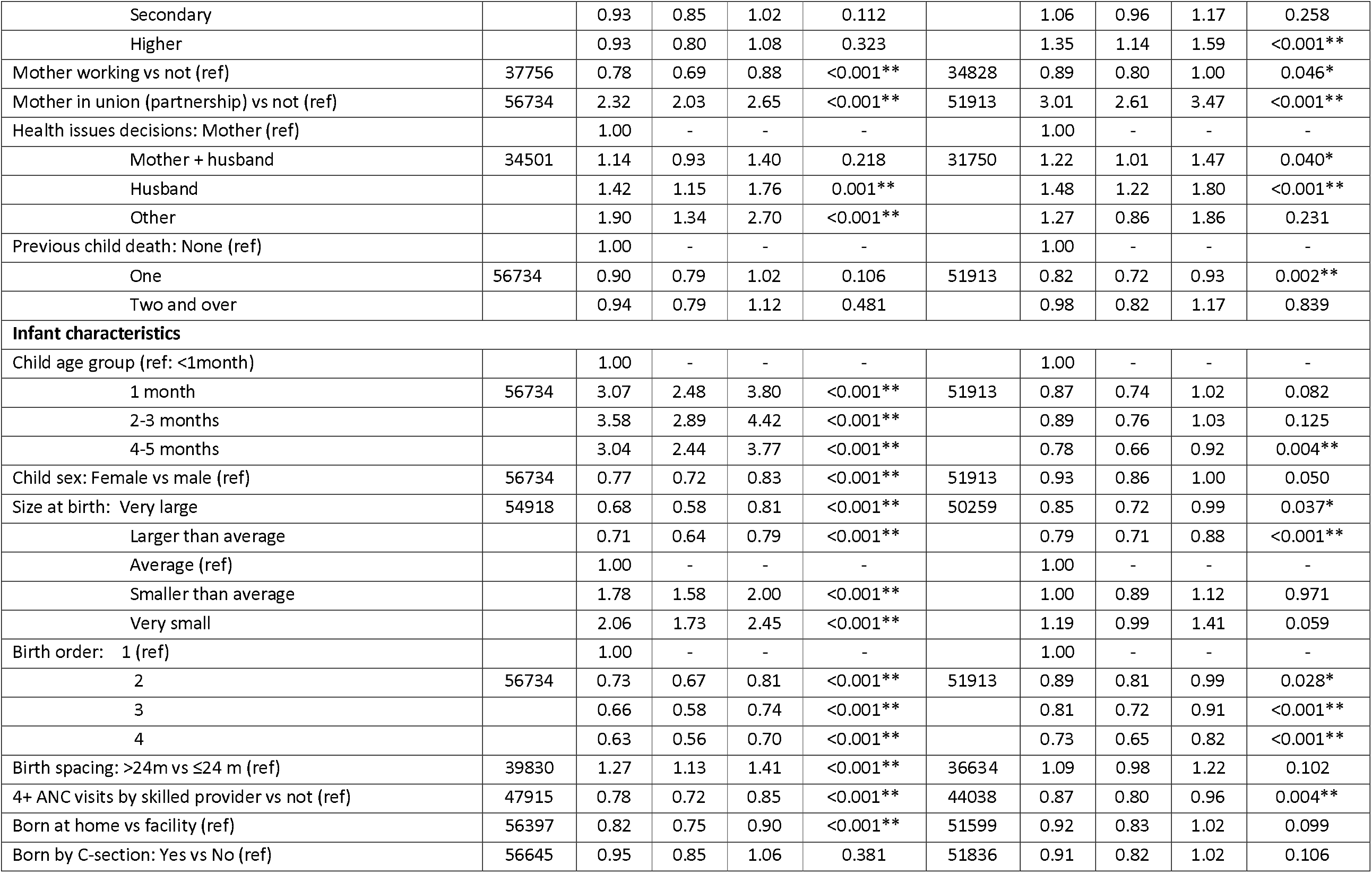
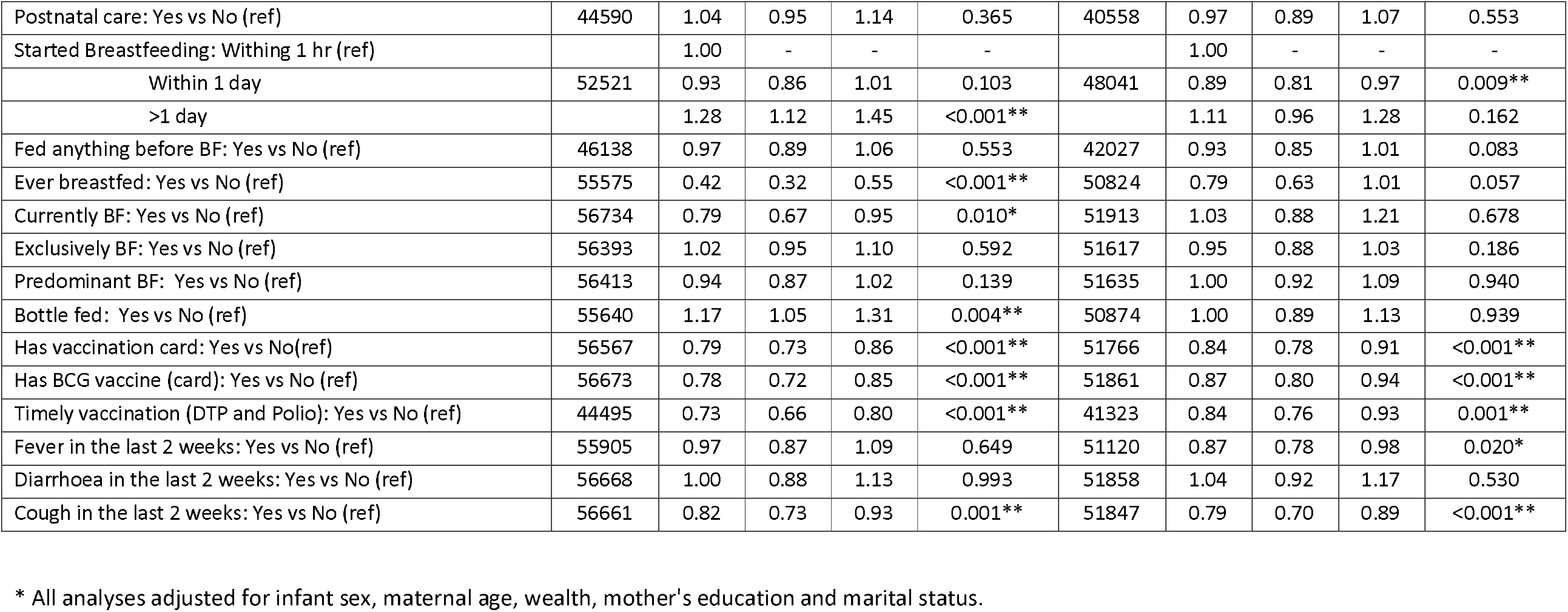
Associations between selected risk factors and underweight and wasted infants u6m.

- maternal age: infants of mothers aged 20-35 less likely to be underweight than those of young mothers aged <20years, OR 0.81 (95% CI: 0.71, 0.92; p<0.001); no such association with wasted infants, OR 0.97 (95% CI: 0.84, 1.13; p=0.725)
- maternal BMI: mothers with low BMI are more likely to have underweight infants, OR 1.65 (95% CI: 1.49, 1.83; p<0.001), and a weaker association, albeit still statistically significant, with wasted infants, OR 1.40 (95% CI: 1.24, 1.57; p<0.001)
- maternal stunting: mothers who are <145cm tall more likely to have underweight infants, OR 2.01 (95% CI: 1.79, 2.25; p<0.001), weaker albeit still statistically significant association with wasted infants, OR 1.27 (95% CI: 1.11, 1.46; p<0.001)
- size at birth: infants reported as having been born “smaller than average” or “very small” are significantly more likely to be underweight compared to those born “average” size, OR 1.78 (95% CI: 1.58, 2.00; p<0.001), OR 2.06 (95% CI: 1.73, 2.45; p<0.001), respectively. In contrast, no such association is observed for wasted infants, OR 1.00 (95% CI: 0.89, 1.12; p=0.971), OR 1.19 (95% CI: 0.99, 1.41; p=0.059), respectively
- receipt of 4+ ANC visits: this is strongly associated with underweight, OR 0.78 (95% CI: 0.72, 0.85; p<0.001), but not as strongly with wasted infants, OR 0.87 (95% CI: 0.80, 0.96; p=0.004)
- ever breastfed: infants u6m ever breastfed were significantly less likely to be underweight, OR 0.42 (95% CI: 0.32, 0.55; p<0.001). They were also less likely to be wasted, but with a weaker OR, 0.79 (95% CI: 0.63, 1.01; p=0.057).

Overall there were six risk factors that were associated with increased odds of both underweight and wasted infants. These comprised having no toilet compared to an improved one, being in the poorest versus middle wealth quintile, maternal underweight compared to normal BMI, maternal stunting, maternal relationship status (being in a union), and the husband making health issue decisions versus the mother. Effect sizes of associations lent towards being stronger with infant underweight for wealth index, maternal underweight and maternal stunting compared to wasted infants, but were weaker for toilet type and being in a union.

There were several risk factors that were only associated with underweight, and not with wasted infants. These included (increasing the odds): being in the second poorest wealth quintile compared to the middle, having an ‘other’ health decision maker compared to the mother, infant age between 1-3 months compared to <1 month, being smaller than average at birth compared to normal, having a birth spacing >24 months versus shorter spacing, starting breastfeeding more than one day after birth compared to within 1 hour, and being bottle fed. Those risk factors decreasing the odds of underweight infants included maternal age >=20 and <35 years compared to <20 years, infant sex (female), being born at home versus in a facility and being ever breast-fed.

There were few risk factors associated only with wasted infants but not with underweight. These were (increasing the odds): having the highest maternal education level compared to none, and having joint health decision making compared to the mother alone; and (decreasing the odds): having one previous child death compared to none, history of fever in the past two weeks, and starting breastfeeding within a day after birth versus within an hour.

One risk factor showed discordant associations. Being aged 4-5 months versus <1 month was strongly associated with increasing the odds of being underweight but was protective against being wasted.

We present the unadjusted univariate results in **Supplementary Table 2**. Adjusting for the *a priori* confounders made minimal difference for the majority of the risk factors, with most relationships maintaining their direction of association and significance. We provide a summary of the significance of associations between all risk factors and both infant outcomes, including both crude and adjusted analyses, in **Supplementary Table 3**.

## DISCUSSION

### Data Quality

Data quality in terms of refusals and missingness was similar for both weight and length but both measures had a high overall percentage of missing data. There were, however, markedly more problem ‘flags’ for the two length-based measures (weight-for-length and length-for-age) than for weight-for-age. This might not be unexpected in a programmatic context given that length is not straightforward to measure. It is, however, very notable in the context of large, well-managed and well-supervised surveys like DHS because surveyors undergo extensive training and have the necessary high-quality equipment to do their jobs well. Such challenges with length measurement and with subsequent length-based indices have been well described previously (15, 16). Our data thus further highlight the risk of low-quality length-based anthropometric measures and indices. Furthermore, any infants with a length <45cm cannot have their WLZ calculated at all (27).

### Burden

The estimated burden of malnutrition among infants u6m in LMICs is large by whichever forms of anthropometric deficit it is assessed. We discuss each form in turn and draw on existing literature where possible for comparison of burden estimates. Though we cannot be certain that the LMIC countries included in our analysis are fully representative all LMIC countries, we do note that the estimated infant u6m population in the 54 countries we examined is 71.4 million out of an estimated total of 122.5 million infants u6m in all LMICs. We therefore cover well over half the estimated total population (58.3%).

Previous estimates for the burden of infant u6m malnutrition focused just on wasted infants. Our work in 2011 (8) suggested approximately 8.5 million infants at the time were wasted. Our current estimates dramatically increase that estimate to 24.5 million wasted infants. This difference is plausible; the previous estimates were based on the 2004 United Nations population database, whereas we used the 2019 database providing estimates for the year 2020. We know that in the 16 years between 2004 and 2020 the world’s population has grown at well over 1.0% per year, and at much higher rates in many LMICs (26). Furthermore, we used 54 DHS surveys in our analyses compared to 21 surveys in the previous paper (8). There are a few comparisons with data other than from DHS surveys. For example, a recent study pooling 18 longitudinal datasets from 10 LMICs found that the highest prevalence of wasted individuals in the cohorts was at birth, which overall was 12% (with a regional maximum of 19% in the South Asian cohorts). Whilst this is lower than our estimated 21.3% from the pooled 54 DHS surveys, the included cohorts in that paper are not nationally representative data (28). The Global Nutrition Report (GNR) updates malnutrition burden estimates annually, pooling data for 0-59 month infants and children. In the 2020 report 7.3% children under 5 years were wasted, corresponding to 49.5 million (29). The GNR does not stratify results for infants u6m, but using our estimates it would suggest anywhere up to half of the GNR estimates of wasted individuals 0-59 months old could be infants u6m. Either the proportion of wasted infants really is this high, or the overall GNR estimates may be underestimating the infant burden; some datasets contributing to the Joint Malnutrition Estimates (on which the GNR is based) exclude infants <6m (30). From our sample of DHS surveys, just considering at the high burden of wasted infants in select populous countries such as India (31.1% wasted; estimated burden of 7.4m wasted infants) and Nigeria (24.6% wasted infants; estimated burden of 1.7m infants) would explain why estimates from our analyses are alarmingly high yet still plausible. Finally, we note that wasting is ideally measured by incidence rather than prevalence since it can be short-lasting and highly seasonal in nature (31). In many countries there are marked peaks around the ‘hungry’ season, when lack of food combines with increased infectious disease transmission: timing of a cross-sectional prevalence survey thus can make all the difference (32). This might account for at least some of the discrepancy we observe.

Our estimates for stunted infants u6m are more consistent with the limited data we find elsewhere. The GNR reported in 2020 that 21.9% infants and children (0-59 months) were stunted, corresponding to 149 million children (29). In the longitudinal analysis of 18 datasets in 10 LMICs, referenced above, the point prevalence of stunted infants at 6 months was 21% (33). In a recent paper combining three longitudinal growth data sets from Malawi, South Africa and Pakistan 20.4% were stunted in the 2 weeks to <3mo age category and 22.6% in the 3 to <6mo category (34). The estimated prevalence of 17.6% stunted infants in our pooled dataset, corresponding to 21.5m stunted infants in all LMICs, therefore seems plausible.

There are no easily accessible global estimates of infants who are underweight (the GNR reports those who are wasted, stunted and overweight), nor for those concurrently wasted and stunted. Regarding the latter, in an analysis of 84 national-level surveys the pooled prevalence of concurrently wasted and stunted children aged 6-59 months was 3.0% (95% CI: 2.97, 3.06) (35). In the above-mentioned paper combining three longitudinal growth data sets from Malawi, South Africa and Pakistan 2.2% were concurrently wasted and stunted in the 2 weeks to <3mo age category and 3.0% in the 3 to <6mo category (34). In the metanalysis of longitudinal datasets from 10 LMICs Mertens *et al*. found that the point prevalence of concurrent WaSt in infants aged 6 months was 1%, increasing to 2% by age 9 months (28).

### Risk factors associated with infant wasting and underweight

Our logistic regression models show that numerous factors are associated with anthropometric deficit in infants u6m. This reflects the long established understanding of malnutrition having diverse causes - immediate, underlying and basic according to the widely-used UNICEF conceptual framework (36). The relative contribution of these causes will of course differ between individual infants u6m. Some causes are also more relevant to infants u6m overall e.g., breastfeeding plays a particularly important role at this age given that their target diet is exclusive breastfeeding (37). Overall, the risk factors in this current analysis are consistent with those found in previous studies (38, 39). Most household, maternal and infant characteristics associated with wasted and underweight infants followed directions of associations found in previous literature (39-44). There were, however, some associations that were not immediately intuitive. Examples of these included some ‘protective’ factors for both wasted and underweight infants, such as having a non-improved toilet compared to an improved one, collecting water outside of the home compared to having water onsite, the maternal relationship being in a union versus not, and the infant having a cough in the last two weeks. These should not be overinterpreted: whenever multiple associations are examined, some will appear statistically significant purely by chance. Though we adjusted for some possible confounders, other confounding may also play a role. Especially in cross-sectional analyses such as ours, we do not claim that any associations, even those with clear mechanistic pathways (e.g., breastfeeding status, socio-economic status, maternal nutritional status), are definitely causal. Stronger study designs such as prospective cohorts and, ideally, intervention trials are needed for such claims. Future research should include developing theoretical frameworks to guide mediation / pathway analysis of direct and indirect moderators of risk, in an attempt to better understand causality of infant malnutrition. What we can say for now, however, is that many factors are likely to play a role in risk of malnutrition, thus multi-faceted solutions will be needed.

With our data being cross-sectional and lacking information on functional outcomes, such as later morbidities and mortality, it means we must be cautious of claims that either weight-for-length (wasting) or weight-for-age (underweight) is a better measure of nutrition-associated risk. We do, however, note that other studies with such functional outcomes, have found that underweight is a better predictor of mortality than wasting (18, 45-47). We do also believe that our data contribute to future policy decisions as to which indicator best reflects nutrition-associated risk. As well as better data quality of underweight, we also draw attention to our observations that more risk factors were associated with underweight than with wasted infants. Of the biologically plausible risk factors that were significantly associated with both outcomes, effect sizes tended to be higher for the underweight model. Remembering that all anthropometric indicators are imperfect proxy measures of malnutrition, this does suggest that underweight reflects malnutrition better than does wasting.

Regardless of the indicators used to describe infant undernutrition, it must be remembered that anthropometric deficit can be a sign of growth deficit but is not a diagnosis in its own right; it is a marker of risk rather than an ultimate outcome in itself (7). Hence, whilst common policy, programme and even research usage refers to infants with anthropometric deficits as “malnourished”, we believe that better future terminology calls them “small and nutritionally-at-risk” (48). Anthropometric deficit might also reflect health and other issues, not just lack of intake of nutrients. Correction of the deficit will not therefore automatically remove associated risk of associated poor outcomes (49). Anthropometry must always be considered alongside wider aetiologic and extenuating factors.

Our analyses particularly highlight the importance of considering maternal factors an integral part of the identification and management of undernutrition in infants. There is no quick fix ‘silver bullet’ than can provide programmatic short-cuts. The prevention and treatment of infant undernutrition needs to consider a package of interventions tackling many possible underlying causes and associated factors (e.g. as in a recent care pathway approach (48)). Our analyses of the potential risk factors for underweight and wasted infants may provide some hypotheses of pathways to examine to ultimately inform whom to target and where to focus contextualised multi-sectoral interventions.

### Limitations

As we emphasise above, our study design comprises of cross-sectional surveys, and therefore associations must not be overinterpreted as necessarily causal. Other limitations are that we included DHS surveys covering a 10-year period during which time risks may have changed. Seasonality affects nutritional outcomes and whilst DHS surveys provide dates of data collection, we do not know how seasonal trends may have affected estimates in each country. DHS also does not have data on nutritional oedema, leading to overall underestimation of the full burden of severe malnutrition (50). Neither did DHS have data on mid-upper arm circumference (MUAC), another common measure of severe malnutrition widely used in older children (51).

Though further confirmatory work is needed, we believe it likely that our estimates of the burden of undernutrition are underestimates. This is mainly because cross-sectional surveys provide information on prevalence but not incidence. For example, in children aged 6-59 months it has been suggested that the true burden of wasting, factoring in incidence, can be anywhere from 1.3 to 30.1 times higher than the prevalence (52). We have, however, assumed our dataset of 54 countries was representative of all LMICs, which may be an over-simplification and may affect numbers downwards as well as upwards. Finally, our risk factor analyses were undertaken to generate hypotheses in exploratory models, hence strict multiple testing was not applied.

## CONCLUSIONS

Malnutrition in infants u6m is a major problem in LMICs. This is true whether assessed by low weight-for-age (underweight), low weight-for-length (wasting) or low length-for-age (stunting). Our data build on other evidence suggesting that underweight may be a better anthropometric case definition than wasting: we show that data quality is better when length is not involved, and underweight is more strongly and consistently associated with biologically plausible risk factors. Future research, ideally from intervention trials, should further explore how best to identify malnourished (small and nutritionally at-risk) infants u6m: measures such as MUAC should be considered alongside weight-for-age and the added value of clinical criteria might also be added. For now, treatment programmes should note that many factors might underlie problems in this age group; services should not only focus on infants themselves but should also care for their mothers and consider wider social circumstances.

## Supporting information

STROBE checklist

COI disclosure

## Data Availability

All data produced are available online. Our data comes from publically-available Demographic and Health Surveys (DHS). Raw survey data is free to download after registration on the DHS website (https://dhsprogram.com/)

## ACKNOWLEDGEMENTS

The authors were able to undertake this work through the generous support of the Eleanor Crook Foundation, and the Department of Foreign Affairs, Ireland. The ideas, opinions and comments included here are entirely the responsibility of the authors and do not necessarily represent or reflect the policies of the donors.

## Author contributions

Paper conceptualisation: MK, MM, SF; data analyses: SF, PJ, EB, CO; wrote the narrative: MK, PJ; all authors contributed to and approved the final draft.

---

**Supplementary Table 1:** List of national surveys included in the analysis, by region

| Country | Phase | Year | N |
| --- | --- | --- | --- |
| ***West and Central Africa*** |  |  |  |
| Burkina Faso | 6 | 2010 | 1,532 |
| Cameroon | 6 | 2011 | 1,194 |
| Chad | 6 | 2014/5 | 1916 |
| Congo | 6 | 2011/2 | 1,002 |
| DRC | 6 | 2013/4 | 2,054 |
| Ghana | 6 | 2014 | 633 |
| Gambia | 6 | 2013 | 994 |
| Guinea | 6 | 2012 | 763 |
| Ivory Coast | 6 | 2011/2 | 836 |
| Liberia | 6 | 2013 | 763 |
| Mali | 6 | 2012/3 | 1,049 |
| Nigeria | 6 | 2013 | 3,151 |
| Niger | 6 | 2012 | 1,364 |
| Senegal | 7 | 2017 | 1,206 |
| Sierra Leone | 6 | 2013 | 1,240 |
| Togo | 6 | 2013/4 | 652 |
| ***East and Southern Africa*** |  |  |  |
| Angola | 7 | 2015/6 | 1,704 |
| Comoros | 6 | 2012 | 353 |
| Burundi | 7 | 2016/7 | 1,290 |
| Ethiopia | 7 | 2016 | 1,137 |
| Gabon | 6 | 2012 | 667 |
| Kenya | 6 | 2014 | 1,891 |
| Lesotho | 6 | 2014 | 347 |
| Malawi | 7 | 2015/6 | 1,748 |
| Mozambique | 6 | 2011 | 1,120 |
| Namibia | 6 | 2013 | 557 |
| Rwanda | 6 | 2014/5 | 727 |
| Sao Tome & Principe | 5 | 2008/9 | 206 |
| Tanzania | 7 | 2015/6 | 1,077 |
| Uganda | 7 | 2016 | 1,590 |
| Zambia | 6 | 2013/4 | 1,242 |
| Zimbabwe | 7 | 2015 | 635 |
| ***Americas*** |  |  |  |
| Bolivia | 5 | 2008 | 819 |
| Dominican Republic | 6 | 2013 | 320 |
| Guatemala | 6 | 2014/5 | 1,206 |
| Guyana | 5 | 2009 | 241 |
| Honduras | 6 | 2011/2 | 1,126 |
| Haiti | 7 | 2016/7 | 747 |
| Peru | 6 | 2012 | 849 |
| **South East Asia & Western Pacific** |  |  |  |
| Bangladesh | 6 | 2014 | 660 |
| Cambodia | 6 | 2014 | 715 |
| India | 6 | 2015/6 | 23,754 |
| Maldives | 5 | 2009 | 422 |
| Myanmar | 7 | 2015/6 | 494 |
| Nepal | 7 | 2016 | 481 |
| Timor-Leste | 7 | 2016 | 787 |
| ***Europe*** |  |  |  |
| Albania | 5 | 2008 /9 | 140 |
| Armenia | 7 | 2015/6 | 182 |
| Kyrgyz Republic | 6 | 2012 | 461 |
| Tajikistan | 6 | 2012 | 435 |
| ***Eastern Mediterranean*** |  |  |  |
| Egypt | 6 | 2014 | 1,540 |
| Jordan | 6 | 2012 | 884 |
| Pakistan | 6 | 2012/3 | 1,155 |
| Yemen | 6 | 2013 | 1,746 |

**Supplementary Table 2:** Crude univariate associations between various risk factors and underweight (WAZ<-2) or wasting (WLZ<-2) in infants u6m

| Risk Factor | All countries |  |  |  |  |  |  |  |  |  |
| --- | --- | --- | --- | --- | --- | --- | --- | --- | --- | --- |
|  | Weight-for-Age Unadjusted |  |  |  |  | Weight-for-Length Unadjusted |  |  |  |  |
|  | N | OR | 95%CI |  | p-value | N | OR | 95%CI |  | p-value |
| Residence: Urban vs rural (ref) | 56734 | 1.17 | 1.07 | 1.27 | <0.001** | 51913 | 1.08 | 0.99 | 1.19 | 0.089 |
| Water source: Improved vs not (ref) | 50526 | 0.73 | 0.66 | 0.80 | <0.001** | 46182 | 0.75 | 0.67 | 0.82 | <0.001** |
| Time to fetch water: On premises (ref) |  | 1.00 | - | - | - |  | 1.00 | - | - | - |
| Source at ≤ 30min | 51942 | 0.86 | 0.79 | 0.93 | <0.001** | 47498 | 0.81 | 0.75 | 0.88 | <0.001** |
| Source at > 30min |  | 0.63 | 0.54 | 0.73 | <0.001** |  | 0.67 | 0.58 | 0.76 | <0.001** |
| Toilet type: Improved (ref) | 52006 | 1.00 | - | - | - | 47577 | 1.00 | - | - | - |
| Non-improved |  | 0.64 | 0.56 | 0.74 | <0.001** |  | 0.60 | 0.53 | 0.69 | <0.001** |
| No toilet |  | 1.45 | 1.34 | 1.57 | <0.001** |  | 1.31 | 1.21 | 1.42 | <0.001** |
| Wealth index: Poorest | 56734 | 1.46 | 1.31 | 1.63 | <0.001** | 51913 | 1.15 | 1.04 | 1.28 | 0.009** |
| Poorer |  | 1.19 | 1.06 | 1.34 | 0.004** |  | 1.03 | 0.92 | 1.15 | 0.584 |
| Middle (ref) |  | 1.00 | - | - | - |  | 1.00 | - | - | - |
| Richer |  | 0.96 | 0.85 | 1.10 | 0.574 |  | 0.98 | 0.86 | 1.12 | 0.780 |
| Richest |  | 0.89 | 0.77 | 1.02 | 0.091 |  | 1.02 | 0.89 | 1.16 | 0.803 |
| Mother's Age: <20 years (ref) |  | 1.00 | - | - | - |  | 1.00 | - | - | - |
| ≥20 & <35 | 56734 | 0.88 | 0.77 | 1.00 | 0.042 | 51913 | 1.09 | 0.94 | 1.27 | 0.252 |
| ≥35 |  | 0.68 | 0.58 | 0.80 | <0.001** |  | 0.78 | 0.65 | 0.94 | 0.008** |
| BMI: <18.5 | 53411 | 1.80 | 1.63 | 1.98 | <0.001** | 48705 | 1.45 | 1.30 | 1.63 | <0.001** |
| ≥18.5 & <25.0 (ref) |  | 1.00 | - | - | - |  | 1.00 | - | - | - |
| ≥25.0 & <30.0 |  | 0.60 | 0.54 | 0.68 | <0.001** |  | 0.81 | 0.72 | 0.90 | <0.001** |
| ≥30.0 |  | 0.55 | 0.44 | 0.68 | <0.001** |  | 0.58 | 0.47 | 0.72 | <0.001** |
| Height: stunted (<145cm) or not (ref) | 53469 | 2.14 | 1.92 | 2.40 | <0.001** | 48756 | 1.30 | 1.14 | 1.48 | <0.001** |
| Mother level of education: No (ref) | 55150 | 1.00 | - | - | - |  | 1.00 | - | - | - |
| Primary |  | 0.59 | 0.53 | 0.65 | <0.001** | 50472 | 0.67 | 0.60 | 0.74 | <0.001** |
| Secondary |  | 0.79 | 0.72 | 0.85 | <0.001** |  | 0.94 | 0.86 | 1.03 | 0.176 |
| Higher |  | 0.73 | 0.64 | 0.84 | <0.001** |  | 1.18 | 1.02 | 1.36 | 0.024* |
| Mother working vs not (ref) | 37756 | 0.76 | 0.68 | 0.85 | <0.001** | 34828 | 0.84 | 0.75 | 0.93 | 0.001** |
| Mother in union vs not (ref) | 56734 | 2.53 | 2.22 | 2.89 | <0.001** | 51913 | 3.22 | 2.80 | 3.71 | <0.001** |
| Health issues decisions: Mother (ref) |  | 1.00 | - | - | - |  | 1.00 | - | - | - |
| Mother + husband | 34501 | 1.23 | 1.01 | 1.51 | 0.039* | 31750 | 1.32 | 1.10 | 1.60 | 0.003** |
| Husband |  | 1.67 | 1.36 | 2.05 | <0.001** |  | 1.64 | 1.35 | 1.99 | <0.001** |
| Other |  | 2.12 | 1.51 | 2.99 | <0.001** |  | 1.44 | 0.99 | 2.11 | 0.057 |
| Child age group (ref: <1month) |  | 1.00 | - | - | - |  | 1.00 | - | - | - |
| 1 month | 56734 | 3.01 | 2.45 | 3.71 | <0.001** | 51913 | 0.89 | 0.76 | 1.04 | 0.139 |
| 2-3 months |  | 3.43 | 2.81 | 4.19 | <0.001** |  | 0.88 | 0.77 | 1.02 | 0.083 |
| 4-5 months |  | 3.26 | 2.66 | 3.98 | <0.001** |  | 0.86 | 0.74 | 1.00 | 0.051 |
| Child sex: Female vs male (ref) | 56734 | 0.78 | 0.73 | 0.84 | <0.001** | 51913 | 0.93 | 0.87 | 1.00 | 0.057 |
| Size at birth: Very large | 54918 | 0.67 | 0.57 | 0.79 | <0.001** | 50259 | 0.79 | 0.68 | 0.93 | 0.003** |
| Larger than average |  | 0.70 | 0.63 | 0.77 | <0.001** |  | 0.75 | 0.67 | 0.83 | <0.001** |
| Average (ref) |  | 1.00 | - | - | - |  | 1.00 | - | - | - |
| Smaller than average |  | 1.76 | 1.58 | 1.96 | <0.001** |  | 0.97 | 0.87 | 1.08 | 0.601 |
| Very small |  | 1.99 | 1.69 | 2.35 | <0.001** |  | 1.16 | 0.98 | 1.39 | 0.093 |
| Birth order: 1 (ref) |  | 1.00 | - | - | - |  | 1.00 | - | - | - |
| 2 | 56734 | 0.77 | 0.71 | 0.85 | <0.001** | 51913 | 0.92 | 0.83 | 1.01 | 0.091 |
| 3 |  | 0.74 | 0.66 | 0.83 | <0.001** |  | 0.84 | 0.75 | 0.94 | 0.003** |
| 4 |  | 0.76 | 0.69 | 0.84 | <0.001** |  | 0.76 | 0.69 | 0.84 | <0.001** |
| Birth spacing: >24m vs ≤24 m (ref) | 39830 | 1.28 | 1.15 | 1.43 | <0.001** | 36634 | 1.11 | 1.00 | 1.23 | 0.045* |
| Previous child death: None (ref) |  | 1.00 | - | - | - |  | 1.00 | - | - | - |
| One | 56734 | 0.96 | 0.85 | 1.08 | 0.479 | 51913 | 0.81 | 0.72 | 0.92 | 0.001** |
| Two and over |  | 1.09 | 0.93 | 1.28 | 0.292 |  | 1.01 | 0.85 | 1.19 | 0.945 |
| 4+ ANC visits by skilled provider vs not (ref) | 47915 | 0.74 | 0.69 | 0.80 | <0.001** | 44038 | 0.87 | 0.80 | 0.95 | 0.001** |
| Born at home vs facility (ref) | 56397 | 0.97 | 0.90 | 1.06 | 0.540 | 51599 | 0.94 | 0.86 | 1.03 | 0.204 |
| Born by C-section: Yes vs No (ref) | 56645 | 0.91 | 0.82 | 1.01 | 0.072 | 51836 | 0.97 | 0.88 | 1.08 | 0.620 |
| Postnatal care: Yes vs No (ref) | 44590 | 0.91 | 0.84 | 0.98 | 0.016* | 40558 | 0.97 | 0.89 | 1.05 | 0.454 |
| Started Breastfeeding: Withing 1 hr (ref) |  | 1.00 | - | - | - |  | 1.00 | - | - | - |
| Within 1 day | 47788 | 1.28 | 1.16 | 1.40 | <0.001** | 43655 | 1.28 | 1.16 | 1.41 | <0.001** |
| >1 day |  | 1.00 | 0.92 | 1.10 | 0.937 |  | 0.97 | 0.88 | 1.06 | 0.497 |
| Fed anything before BF: Yes vs No (ref) | 46138 | 1.01 | 0.93 | 1.09 | 0.891 | 42027 | 0.93 | 0.86 | 1.02 | 0.120 |
| Ever breastfed: Yes vs No (ref) | 55575 | 0.45 | 0.35 | 0.58 | <0.001** | 50824 | 0.78 | 0.62 | 0.99 | 0.044* |
| Currently BF: Yes vs No (ref) | 56734 | 0.86 | 0.73 | 1.01 | 0.073 | 51913 | 1.08 | 0.92 | 1.26 | 0.359 |
| Exclusively BF: Yes vs No (ref) | 56393 | 0.94 | 0.88 | 1.01 | 0.107 | 51617 | 0.98 | 0.91 | 1.05 | 0.574 |
| Predominant BF: Yes vs No (ref) | 56413 | 0.94 | 0.87 | 1.02 | 0.118 | 51635 | 1.05 | 0.97 | 1.14 | 0.204 |
| Bottle fed: Yes vs No (ref) | 55640 | 1.13 | 1.02 | 1.25 | 0.017* | 50874 | 0.96 | 0.86 | 1.07 | 0.486 |
| Has vaccination card: Yes vs No(ref) | 56567 | 0.78 | 0.72 | 0.84 | <0.001** | 51766 | 0.80 | 0.75 | 0.86 | <0.001** |
| Has BCG vaccine (card): Yes vs No (ref) | 56673 | 0.77 | 0.72 | 0.83 | <0.001** | 51861 | 0.82 | 0.77 | 0.89 | <0.001** |
| Timely vaccination (DTP and Polio): Yes vs No (ref) | 44495 | 0.62 | 0.57 | 0.68 | <0.001** | 41323 | 0.82 | 0.75 | 0.90 | <0.001** |
| Fever: Yes vs No (ref) | 55905 | 1.04 | 0.94 | 1.16 | 0.449 | 51120 | 0.86 | 0.77 | 0.96 | 0.006** |
| Diarrhoea: Yes vs No (ref) | 56668 | 1.08 | 0.96 | 1.21 | 0.179 | 51858 | 1.03 | 0.92 | 1.16 | 0.621 |
| Cough: Yes vs No (ref) | 56661 | 0.84 | 0.75 | 0.94 | 0.002** | 51847 | 0.73 | 0.65 | 0.83 | <0.001** |

**Supplementary Table 3:** Summary of significance of associations between risk factors and outcomes in crude analyses and adjusted analyses

|  | All |  |  |  |
| --- | --- | --- | --- | --- |
|  | WAZ | WLZ | WAZa<sup>Ⓜ</sup> | WLZa<sup>Ⓜ</sup> |
| Residence: Urban vs rural (ref) | ** | - | - | - |
| Water source: Improved vs not (ref) | ** | ** | ** | ** |
| Time to fetch water: On premises (ref) | ** | ** | ** | ** |
| Toilet type: Improved (ref) | ** | ** | ** | ** |
| Wealth index: Middle(ref) | ** | ** | ** | ** |
| Mother's Age: <20 years (ref) | ** | ** | ** | ** |
| BMI: ≥18.5 & <25.0 (ref) | ** | ** | ** | ** |
| Height: stunted (<145cm) or not (ref) | ** | ** | ** | ** |
| Mother level of education: No (ref) | ** | ** | ** | ** |
| Mother working vs not (ref) | ** | ** | ** | * |
| Mother in union vs not (ref) | ** | ** | ** | ** |
| Health issues decisions: Mother (ref) | ** | ** | ** | ** |
| Child age group (ref: <1month) | ** | - | ** | ** |
| Child sex: Female vs male (ref) | ** | - | ** | - |
| Size at birth: Average (ref) | ** | ** | ** | ** |
| Birth order: 1 (ref) | ** | ** | ** | ** |
| Birth spacing: >24m vs ≤24 m (ref) | ** | * | ** | - |
| Previous child death: None | - | ** | - | ** |
| 4+ ANC visits by skilled provider vs not (ref) | ** | ** | ** | ** |
| Born at home vs facility (ref) | - | - | ** | - |
| Born by C-section: Yes vs No (ref) | - | - | - | - |
| Postnatal care: Yes vs No (ref) | * | - | - | - |
| Started Breastfeeding: Withing 1 hr (ref) | ** | ** | ** | ** |
| Fed anything before BF: Yes vs No (ref) | - | - | - | - |
| Ever breastfed: Yes vs No (ref) | ** | * | ** | - |
| Currently BF: Yes vs No (ref) | - | - | * | - |
| Exclusively BF: Yes vs No (ref) | - | - | - | - |
| Predominant BF: Yes vs No (ref) | - | - | - | - |
| Bottle fed: Yes vs No (ref) | * | - | ** | - |
| Has vaccination card: Yes vs No(ref) | ** | ** | ** | ** |
| Has BCG vaccine (card): Yes vs No (ref) | ** | ** | ** | ** |
| Timely vaccination (DTP and Polio): Yes vs No (ref) | ** | ** | ** | ** |
| Fever: Yes vs No (ref) | - | ** | - | * |
| Diarrhoea: Yes vs No (ref) | - | - | - | - |
| Cough: Yes vs No (ref) | ** | ** | ** | ** |

\*p<0.05
\*\*p<0.001
<sup>Ⓜ</sup>adjusted models

